# Multi-molecular phenotyping in a self-sampling population

**DOI:** 10.1101/2025.01.14.25320405

**Authors:** Leo Dahl, Annika Bendes, María Bueno Álvez, Vincent Albrecht, Hooman Aghelpasand, Sophia Björkander, Simon Kebede Merid, Anja Mezger, Max Käller, Claudia Fredolini, Åsa Torinsson Naluai, Olof Beck, Erik Melén, Stefan Bauer, Magnus Gisslén, Niclas Roxhed, Jochen M Schwenk

## Abstract

The recent COVID-19 pandemic has posed major health challenges. To expand our molecular understanding beyond those seeking medical care, we conducted a cross-sectional survey targeting random residents from Sweden’s two largest cities.

Two thousand participants were invited during 2021 to self-sample dried blood spots (DBS) and provide health information. DBS samples were analysed for multiple anti-SARS-CoV-2 antibodies (Abs), auto-reactive antibodies (AAbs) against 23 interferons (IFNs), and for 502 circulating immune-related proteins. Data-driven methodologies characterised the DBS phenotypes in response to infections and vaccination.

We received DBS samples from 437 (22%) volunteers aged 18-69, 60% females, and 50% per city. Multi-analyte serology distinguished self-reported infections (26%) and vaccination (40%), revealing a time-dependent discrepancy between reported and measured immunity. Anti-IFN Abs were detected in 21% of the donors and more frequent for type 1 IFNs alongside a natural infection. Integrating proteomics with antibody data provided additional insights into processes of cell-mediated immunity. Proteomics-centric analysis identified 24% of the participants to deviate in their phenotypes due to infection, immunity, respiratory distress, and age-related traits.

Multi-molecular analysis of layperson samples uncovered heterogeneity and diversity of immunity and health phenotypes, complementing clinical investigations.

## Introduction

Since the beginning of the global COVID-19 pandemic, the consequences of SARS-CoV-2 virus infections, disease severity, and longer-term health consequences continue to be studied extensively. A logical early focus was to ensure the well-being of those requiring clinical support due to severe illness or those at risk of more profound health effects. With the end of the pandemic, those suffering from post-acute sequelae of COVID-19 (PASC) have received more attention. It is estimated that > 750 million people have been infected by the virus globally [1] (https://covid19.who.int). In Sweden, more than 97% of the population was seropositive for SARS-CoV-2 spike in October 2024 [2], due to previous infections and vaccinations. The seroprevalence was also over 95% in children under 14 years of age - a group not prioritised for vaccination in Sweden, with only 1% receiving the vaccine. This indicates a very high prevalence of prior infections within the population. Estimations based on antibody (Ab) tests conducted during 2021 showed that > 30% of the Swedish population had already been infected by that time [3]. At the same time, only a small fraction of those infected required hospitalisation or sought medical care. This leaves a gap in understanding of those who never required medical care or enrolled in clinical studies due to health interests, meaning that most of the public did not get involved. A general hurdle was reaching those not actively joining research studies. Hence, there is a lack of molecular data for studying the heterogeneity of population health phenotypes and in relation to combinations of infections and vaccinations.

It is here where self-sampling devices have become a valuable tool to bridge between a person’s home and advanced laboratory methods. With blood being the most commonly used specimen in clinical diagnostics, newer devices have been developed that collect quantitative volumes of blood from a simple finger prick [4]. Once collected, the dried blood specimen can be shipped by regular mail, circumventing the need for a donor to book an appointment, travel to a health care center, and meet a staff member trained in venous blood draws. In line with these developments and use for COVID-19 antibody testing [5], [6], [7], omics technologies have adapted their workflows to meet the requirements of small sample volumes and maintain analytical precision, robustness, and depth. Among these approaches, the study of circulating proteins utilising mass spectrometry or affinity-based assays can offer valuable insights into the current health status of a person [8].

It is well-established that infections by SARS-CoV-2 affect the circulating proteome [9], [10]. In particular, blood proteins related to inflammatory response, manifested in cytokine storms [11] and immune cell receptor activation [12], revealed wide-ranging health consequences. Multi-analyte approaches have been established to study the serological response to infection and vaccination [13]. In addition, the repertoire of anti-cytokine autoantibodies has also been shown to be affected by infection [14], and auto-reactive antibodies (AAbs) targeting a wide range of proteins [15] have been linked to COVID-19, some emerging on infection [16]. Notably, AAbs against type I interferons (IFN-I) have been linked to COVID-19 severity [17], particularly in males [18]. Further evidence suggested that anti-IFN AAbs were associated with SARS-CoV-2 infections rather than vaccine response [19]. Due to the diverse mechanisms of a proteome-centric response, highly discriminative treatment is needed to differentiate infections by different virus variants and healthy controls [20].

Here, we used quantitative dried blood spots (qDBS) collected in randomly invited individuals from Stockholm and Gothenburg in Sweden to elucidate the molecular consequences of SARS-CoV-2 infection and vaccination. Together with self-reported data on health-related aspects, we analysed the DBS samples for anti-viral, auto-reactive, and proteomic responses using previously established multiplexed assay workflows [6], [21]. The study confirmed distinct immune responses to infection and vaccination and their influence on circulating inflammatory proteins. We observed differences between reported and molecular immune states and determined the heterogeneity among population phenotypes. The study confirms the utility of self-sampling to provide molecular information to supplement perceived health information.

## Methods and materials

### Samples

Samples were obtained from individuals randomly chosen from the population registers of metropolitan Stockholm and Gothenburg by mailing home-sampling kits (MM18-01-001, Capitainer AB, Sweden) to 1000 individuals of ages 18-69 years in each city as previously described [6], [21]. Anonymous volunteering participants sampled capillary blood per instructions by pricking a finger and collecting droplets on a quantitative DBS sampling card. Participants also filled out a questionnaire with health-related questions and a consent form. Sampling cards, questionnaires, and consent forms were returned by regular mail, and the cards were stored at room temperature until extraction. The study was approved by the regional ethical board (EPN Stockholm, Dnr 2015/867-31/1) and the Swedish Ethical Authority (EPM, Dnr 2021-01106 and Dnr 2023-03016-02).

### Sample preparation

The DBS cards were eluted to obtain samples for analysis as previously described [6], [21]. In brief, the cards were first inactivated by heating in an oven (UN55m, Memmert GmbH) at 56°C for 60 minutes. The inactivated discs containing blood were placed in a flat-bottom 96-well plate (#734-2327, VWR) and treated with 100 µL PBS with 0.05% Tween20 (#97062-332, VWR) and protease inhibitor cocktail (#04693116001, Roche). The plates were shaken gently at 170 rpm at room temperature for 60 min and then centrifuged (2095 rcf, Allegra X-12R, Beckman Coulter Inc) at 3000 rpm for 3 min before collecting 70 µL of supernatant in a PCR plate (#732-4828, VWR). The eluates were stored at -20°C until analysis.

### Serological assays

Serological assays were performed on samples from 437 participants who successfully completed both the blood spot collection and questionnaire. IgG antibodies against the nucleocapsid (N), spike (S and RBD) proteins of the SARS-CoV-2 virus, as well as against several human interferons, were measured using suspension bead arrays (SBAs). The antigens that were used are shown in Table 1. The assays were performed as previously described [6], [21]. In brief, target proteins were covalently linked to colour-coded magnetic beads (MagPlex, Luminex Corp.) with NHS/EDC coupling, and the beads were combined to form an antigen bead array. DBS eluates were diluted 2.5x with assay buffer (PBST with 3% BSA and 5% milk powder), and 35 µL of each was combined with a 5 µL antigen bead array and incubated at room temperature for 60 min. The beads were analysed using a Luminex FlexMap 3D instrument (Luminex Corp) with anti-human IgG-R-PE (Jackson ImmunoResearch) for detection. Measurements were reported as median fluorescence intensity (MFI) values per antigen and sample, where each data point had at least 32 events per bead ID.

**Table 1.** Self-reported demographics and symptoms. Self-reported sample demographics by sampling region and in total.

|  | Gothenburg | Stockholm | Overall |
| --- | --- | --- | --- |
| **Participants** | 220 (50.3%) | 217 (49.7%) | 437 (100%) |
| **Age group** |  |  |  |
| 18-29 | 43 (19.5%) | 33 (15.2%) | 76 (17.4%) |
| 30-39 | 42 (19.1%) | 48 (22.1%) | 90 (20.6%) |
| 40-49 | 53 (24.1%) | 40 (18.4%) | 93 (21.3%) |
| 50-59 | 48 (21.8%) | 60 (27.6%) | 108 (24.7%) |
| 60-69 | 34 (15.5%) | 36 (16.6%) | 70 (16.0%) |
| **Sex** |  |  |  |
| Female | 120 (54.5%) | 140 (64.5%) | 260 (59.5%) |
| Male | 99 (45.0%) | 76 (35.0%) | 175 (40.0%) |
| Missing | 1 (0.5%) | 1 (0.5%) | 2 (0.5%) |
| **Symptoms (loss of)** |  |  |  |
| None | 178 (80.9%) | 170 (78.3%) | 348 (79.6%) |
| Smell | 9 (4.1%) | 9 (4.1%) | 18 (4.1%) |
| Taste | 5 (2.3%) | 3 (1.4%) | 8 (1.8%) |
| Smell + taste | 28 (12.7%) | 35 (16.1%) | 63 (14.4%) |
| **Vaccination** |  |  |  |
| Not vaccinated | 145 (65.9%) | 123 (56.7%) | 268 (61.3%) |
| 1 dose | 57 (25.9%) | 76 (35.0%) | 133 (30.4%) |
| 2 doses | 18 (8.2%) | 17 (7.8%) | 35 (8.0%) |
| Missing | 0 (0%) | 1 (0.5%) | 1 (0.2%) |
| **Infection** |  |  |  |
| No | 169 (76.8%) | 154 (71.0%) | 323 (73.9%) |
| Yes | 51 (23.2%) | 63 (29.0%) | 114 (26.1%) |

### Autoreactive antibody assays

In addition to the COVID-19 antigens, we included 22 human interferons (IFNs) in the bead mixture of the SBAs, see Table S1. The IFNs were produced as described elsewhere [22]. Protein coupling, analysis, and readout were conducted as described for SARS-CoV-2 proteins above. Since IFN assays report the levels of auto-reactive antibodies (AAbs) against different interferons, we conducted the data analysis separately. The eluates were stored at -80°C until further analysis.

### Proteomics assays

To measure a large number of low-abundant and disease-relevant circulating inflammatory proteins for comparison with the serology, samples were sent to SciLifeLab’s Affinity Proteomics Unit (Uppsala, Sweden) for the proximity extension assay (PEA) using Olink Explore [23], and to Alamar Biosciences Inc. (Fremont, California, USA) for the NULISAseq assay [24].

### Proximity extension assay

The samples were processed with Olink Explore 384 Inflammation panel (Olink Proteomics AB, Uppsala, Sweden), which is based on Proximity Extension Assay (PEA) [23] and allows for simultaneous detection of up to 368 proteins using 1 µL of DBS eluate. In brief, two matched antibodies labeled with unique complementary oligonucleotides bind to a target protein in solution, which brings the oligonucleotides in proximity and allows them to hybridise and extend to form a double-stranded DNA barcode unique to the target protein. The subsequent library preparation adds sample indices and sequencing adapters, enabling multiplexing and readout with next-generation sequencing. The manufacturer’s instructions were followed, except that Olink and Science for Life Laboratory in Stockholm collaborated to enable the Agilent Bravo (Agilent Technologies, Santa Clara, CA, USA) automated liquid handler to be used in the workflow. The libraries were sequenced on a NovaSeq 6000 (Illumina, San Diego, CA, USA) at the Science for Life Laboratory in Uppsala, Sweden, according to Olink instructions. The data are reported as Normalised Protein eXpression (NPX) values, semi-quantitative arbitrary units (AU) on a log2 scale.

### NULISAseq assay

NULISAseq assays were performed at Alamar Biosciences, Fremont, USA, as described previously [24]. Briefly, DBS samples were shipped on dry ice, stored at -80°C, thawed on ice and centrifuged at 10,000g for 10 mins. Then, 10 µL supernatant samples were plated in 96-well plates and analysed with Alamar’s Inflammation Panel 250, targeting mostly inflammation and immune response-related cytokines and chemokines. A Hamilton-based automation instrument was used to perform the NULISAseq workflow, starting with immunocomplex formation with DNA-barcoded capture and detection antibodies, followed by capturing and washing the immunocomplexes on paramagnetic oligo-dT beads, then releasing the immunocomplexes into a low-salt buffer, which were captured and washed on streptavidin beads. Finally, the proximal ends of the DNA strands on each immunocomplex were ligated to generate a DNA reporter molecule containing target-specific and sample-specific barcodes. DNA reporter molecules were pooled and amplified by PCR, purified, and sequenced on Illumina NextSeq 2000.

Sequencing data were processed using the NULISAseq algorithm (Alamar Biosciences). The sample-(SMI) and target-specific (TMI) barcodes were quantified, and up to two mismatching bases or one indel and one mismatch were allowed. Intraplate normalisation was performed by dividing the target counts for each sample well by that well’s internal control counts. Inter-plate normalisation was then performed using inter-plate control (IPC) normalisation, wherein counts were divided by target-specific medians of the three IPC wells on that plate. Data were then rescaled, add 1 and log2 transformed to obtain NULISA Protein Quantification (NPQ) units for downstream statistical analysis.

### Data analysis

Data processing, analysis and visualisation was performed with the R (version 4.3.2) [25] and Julia (version 1.7.2) [26] programming languages and the tidyverse (version 2.0.0) [27], targets (version 1.4.1) [28], and tarchetypes (version 0.7.12) [29] R packages. Plots were created using either the ggplot2 package (version 3.5.1) [30] together with the patchwork (version 1.3.0.9000) [31], RColorBrewer (version 1.1.3) [32], ggh4x (version 0.2.8) [33], ggnewscale (version 0.4.10) [34], ggbeeswarm (version 0.7.2) [35], ggrepel (version 0.9.5) [36], ggsignif (version 0.6.4) [37], ggbiplot (version 0.6.2) [38], GGally (version 2.2.1) [39] and ggvenn (version 0.1.10) packages [40], or the ComplexHeatmap (version 2.18.0) [41] and circlize (version 0.4.15) [42] packages. The table1 package (version 1.4.3) [43] was used to generate some of the tables.

### Serological data analysis

Data from the serological assay were normalised to remove correlation with negative control samples using a mixed model programmed in Julia. The model was fit on log2-transformed data and consisted of a linear model and a uniform distribution model to account for distributions where most samples were at low baseline levels while a few samples were at higher positive levels. The resulting relative log2 values were scaled and centered per protein to put them on the same scale. An overview of the antibody data distribution is given in supplementary file 1.

Seropositivity of each antibody was determined using a population-based threshold at 6 SD (for anti-SARS-CoV-2 Abs) or 12 SD (for IFN AAbs) of the estimated negative proportion (uninfected individuals for N and IFNs, uninfected and unvaccinated individuals for S, RBD) above the gaussian population peak as described previously [6]. Attained seropositivity classifications for IFN AAbs were associated with questionnaire data and anti-SARS-CoV-2 Ab classifications using the Fisher exact test.

Hierarchical clustering was used on the N, S, and RBD MFI data to stratify samples based on serology. The resulting clustering was cut into four seroclusters that were compared to vaccination and infection status from the questionnaire. Mismatch in questionnaire data and seroclusters regarding vaccination was tested using the Fisher exact test. The clustering was evaluated with Lasso regularised logistic or multinomial regression using the collection of functions in the tidymodels package (version 1.1.1.) [44] to predict questionnaire traits and seroclusters using anti-SARS-CoV-2 Ab levels. 70% of the samples were used as a training set and the remaining 30% as a hold-out test set. The Lasso regression lambda hyperparameter was tuned using five-fold cross-validation on the training set.

### Proteomics data analysis

Normalised protein measurements were received from Olink and Alamar. To reduce sample-to-sample variation stemming from sample handling, protein measurements were normalised with the ProtPQN method using the ProtPQN package (version 1.0.1) [45]. Deviating samples were detected by plotting sample medians and interquartile ranges (IQRs). Samples falling outside of 3 SDs of either axis were excluded from downstream analysis. Confounding from sex differences was removed using a linear model. An overview of the proteomics data distribution is given in supplementary file 2. The data were plotted using the principal component analysis (PCA) dimensionality reduction technique to visually check for trends in the data.

To compare circulating proteins with serology, proteins were associated with seropositivity classifications of anti-SARS-CoV-2 Abs and IFN AAbs using logistic regression with the seropositivity as outcome. The proteins were also associated with the seroclusters using the Kruskal-Wallis test and, if FDR < 0.05, the Wilcoxon rank-sum test for pair-wise comparisons. Furthermore, to investigate the predictive power of proteins on age, sex, region, and immune status defined by the questionnaire and the seroclustering, Lasso regularised logistic or multinomial regression was performed similarly to the serology.

Clustering was performed on the proteomics data to find protein-centric sample groupings similarly to the serology data. Unlike the low-dimensional serology data analysis, the proteomics data dimensions were reduced using weighted gene correlation network analysis (WGCNA) using the WGCNA R package (version 1.72-5) [46]. The pickSoftThreshold function was used to pick the power and WGCNA was performed using the blockwiseModules function with default parameters. The resulting protein module eigengenes (MEs, apart from the grey module containing non-correlated proteins) were used to cluster the samples with hierarchical clustering where the gap statistic was used to determine the optimal number of clusters (cluster package version 2.1.4 [47]). The cluster stability was evaluated by calculating the mean Jaccard index for each cluster with the clusterboot function of the fpc package (version 2.2-11) [48]. Any clusters with a mean Jaccard index below 0.5 or a size below 10 were aggregated into a noise cluster. The resulting clusters, henceforth called proteotypes, were numbered from 0 (noise) to 5.

Associations between WGCNA protein modules and population traits and diseases were investigated by integrating previously reported UK Biobank associations [49]. The proportions of significantly associated proteins for each module and trait were compared to the proportions in the rest of the samples using the Fisher exact test. For HPA, the proportions of proteins from each module were plotted for each immune cell type.

The modules’ links to immunity were characterised by studying immune cell type enrichment annotation from the Human Protein Atlas (HPA) and Gene Ontology Biological Process (GO-BP) term enrichment using the STRING database (version 12.0) [50]. For HPA, the proportions of proteins from each module were plotted for each immune cell type. In the comparative GO-BP enrichment analysis, the whole genome was used as background to identify differences between the modules.

The WGCNA module differences between proteotypes were visualised by calculating the differences in ME means for each proteotype against the remaining proteotypes. The 95% confidence intervals were also calculated using the t.test R function to quantify the confidences of the differences. Differences in module distributions across seroclusters, CRP level classes (determined using the population threshold described above with 3 and 6 SD), and vaccine doses were compared using the Fisher exact test.

## Results

We have studied the molecular impact of SARS-CoV-2 infections and vaccination in citizens of Sweden’s two largest metropolitan areas. During the summer of 2021, we invited randomly selected citizens to provide DBS samples and answer a questionnaire. The goal was to describe how COVID-19 influenced the serological and proteomic phenotypes in non-hospitalised participants from the public.

### Study design and demographics

Home sampling kits and questionnaires were sent out to 2000 random inhabitants of Gothenburg and Stockholm in April 2021 to obtain samples from a widespread population in Sweden. We invited 1000 citizens of each city, targeting those aged 18 to 69 years and 50% split between sexes (Fig 1a-b). We received 478 (23.9%) kits by postal mail, and after removing contributions with incomplete sampling, missing questionnaires, and consent forms, 437 (21.9%) donors were included for further analyses.

**Fig. 1:**
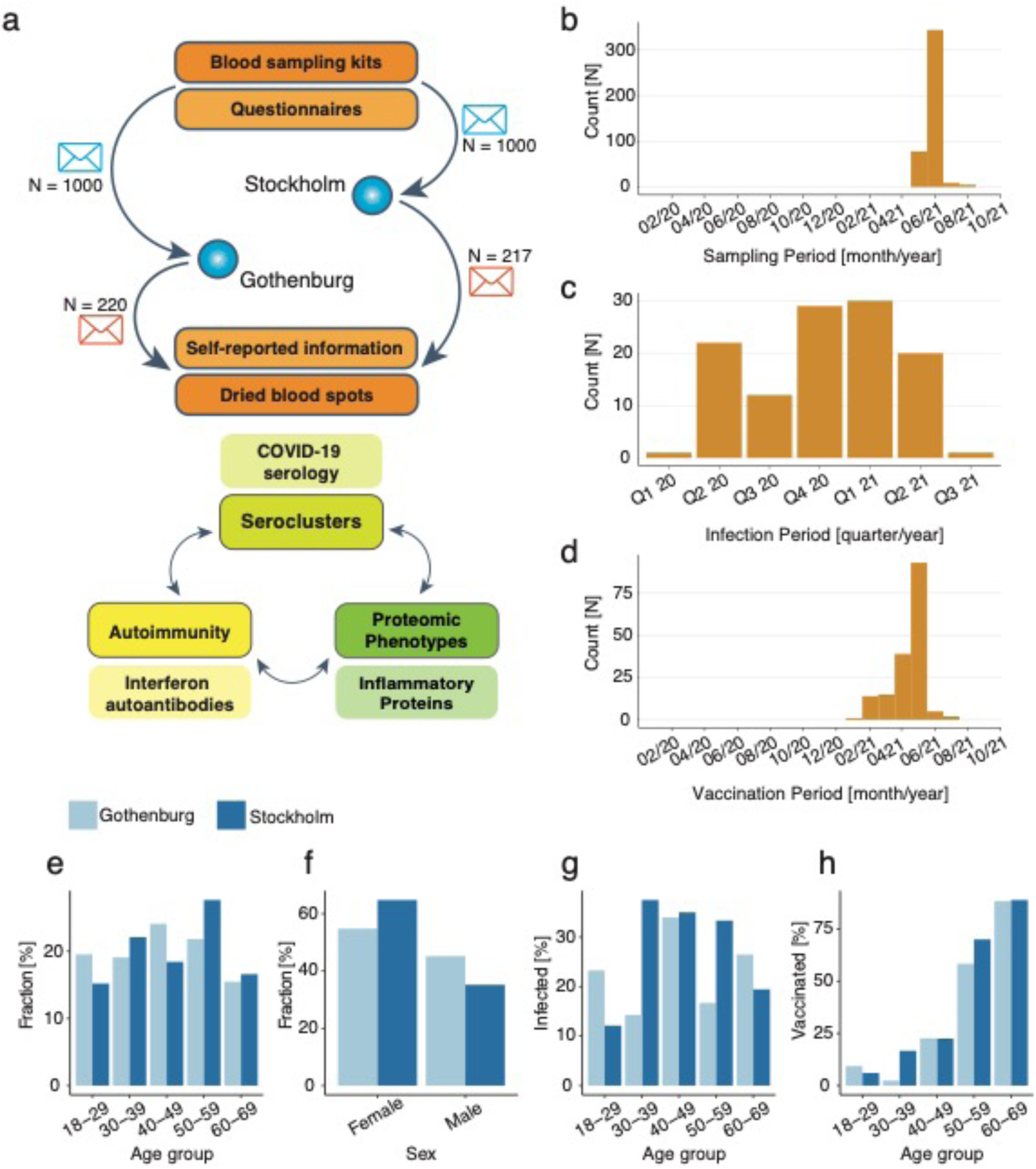
Study design and cohort demographics. **(a)** DBS home sampling kits were sent to 2000 randomly selected households in Stockholm and Gothenburg. Of the returned kits, 437 were used for downstream analysis. **(b)** Samples were taken between May and August 2021, peaking around June. **(c)** Self-reported data showed that infection by SARS-CoV-2 was spread out between the beginning of 2020 and Q3 of 2022, **(d)** while vaccination was concentrated around January 2021 to July 2021. **(e)** The age of the participants ranged between 18 and 69 years, with fewer participants being in the youngest and oldest age groups. **(f)** In both regions, women were more likely to return samples than men. **(g), (h)** Young participants were less likely to have been infected and vaccinated, with vaccination being more common with increasing age.

Of the 437 analysed samples, 220 (50.3%) were from Gothenburg and 217 (49.7%) were from Stockholm (Table 1). In both regions, more women participated than men (Fig 1f), and compared to population statistics, our study contained more donors aged 40-59 (Fig 1e). Overall, 38.4% of individuals reported being vaccinated at least once, and 26.1% reported a COVID-19 infection (confirmed by a diagnosis, a PCR test or an Ab test) before sampling (Fig 1c, 1g). Vaccination rate followed the rollout strategy (Fig 1d, 1h), with individuals aged 50-59 and 60-69 having a higher rate of vaccination (16.0% and 14.2%, respectively) than individuals in the age groups 18-29, 30-39, and 40-49 (1.37%, 2.06%, and 4.81%, respectively).

This data aligns well with population statistics in Sweden, suggesting that already > 30 % of the public had been exposed to the virus earlier in 2021 [3], and that nearly 40% had received a vaccine via the national program by early June [51].

### Multi-analyte serology

Based on previous work [6], [21], we performed multi-analyte serology of circulating human IgG antibodies in DBS samples to determine who developed an immune response against two major SARS-CoV-2 antigens. We used a data-driven cut-off without community standards for DBS samples to classify samples as seropositive. To increase the certainty of our observations, multiple proteins for spike (S) and nucleocapsid (N) were included alongside the receptor binding domain (RBD) of S. As shown in Table 2, up to 50% of the samples were deemed seropositive for anti-S IgG (denoted S+) and around 20% of the samples contained IgG against N (denoted N+).

**Table 2:** Multi-analyte SARS-CoV-2 serology. SARS-CoV-2 antigens, their sources, and the percent of the samples determined to be seropositive, alongside 95% confidence intervals (CI).

| Acronym | SARS-CoV-2 Protein | Source | % positive | 95% CI |
| --- | --- | --- | --- | --- |
| **S1** | Spike, S1 domain | KTH | 45.3 | 40.6 – 50.0 |
| **S1S2** | Spike, foldon | KTH | 50.3 | 45.6 – 55.0 |
| **RBD** | Spike, receptor binding domain | KTH | 31.1 | 26.8 – 35.4 |
| **Na** | Nucleocapsid | Acro | 20.6 | 16.8 – 24.4 |
| **Nc** | Nucleocapsid, C-terminal | KTH | 19.0 | 15.3 – 22.7 |

It is well-established that infection with SARS-CoV-2 triggers production of both anti-S and anti-N (S+N+) antibodies, while vaccination against the virus leads to production of only anti-S (S+N-) antibodies. Self-reported data suggested that 38% of all donors were vaccinated, 26% experienced an infection, and 9.4% reported both. As shown in Fig. 2a-c for both collection regions, elevated levels of anti-S and anti-RBD were found in samples of donors who reported a natural infection and vaccination, while anti-N levels were elevated in infected participants. This explains the higher percentage of samples classified as S+ compared to N+. We also observed that 23% of infected or vaccinated participants had lower anti-S and anti-RBD levels than expected from the questionnaire and that 62% of donors who reported being infected and 2.2% of donors reporting no infection were N+. Fig 2d shows the relative antibody levels against the Epstein Barr virus (EBV) antigen EBNA1. Since ∼ 90% of the general public has been exposed to EBV [52], it served as a control for the immune response against SARS-CoV-2.

**Fig 2:**
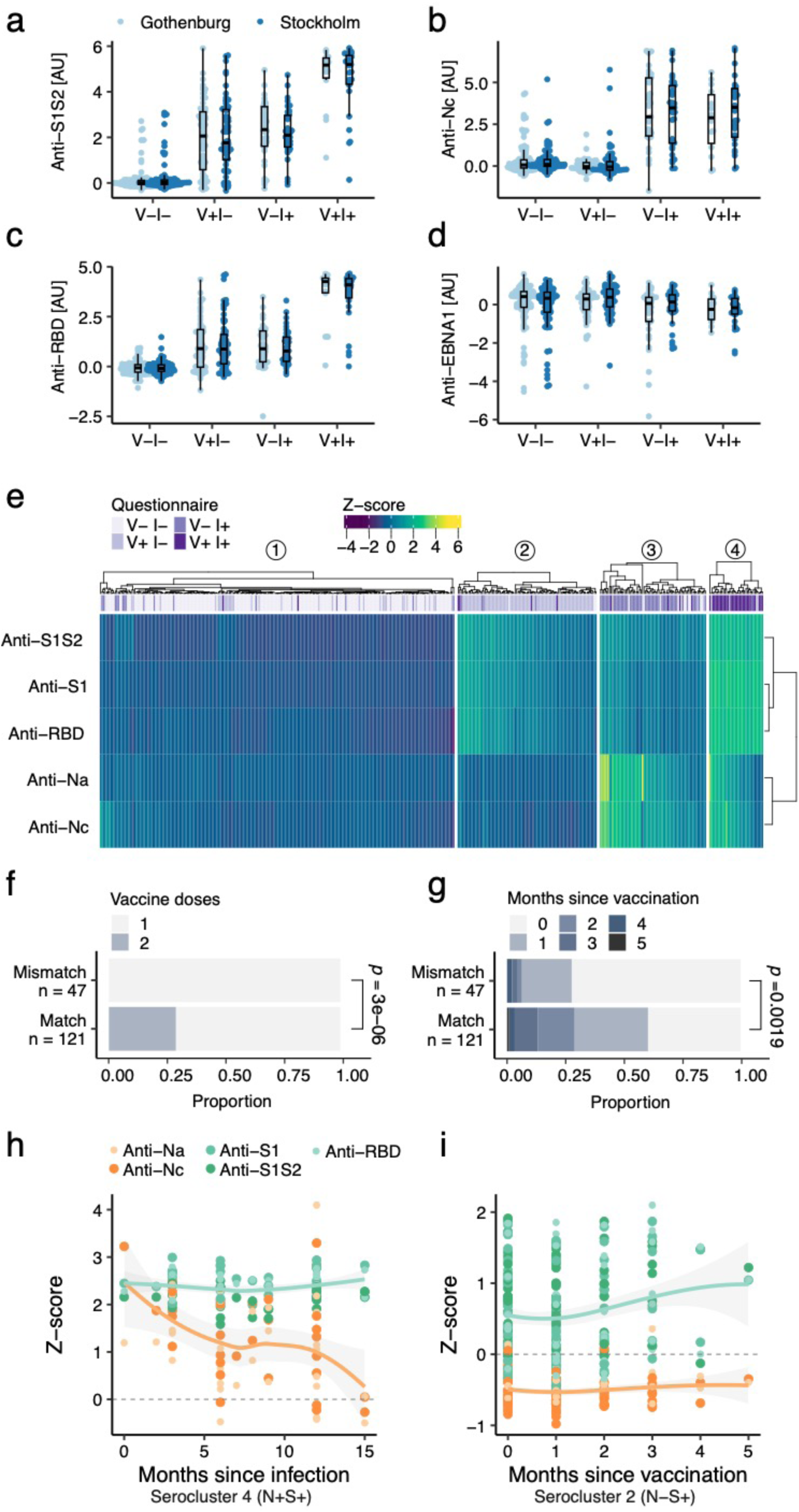
Serological characterisation. (a-c) Levels of Abs targeting SARS-CoV-2 spike protein were highest in individuals who were both infected and vaccinated (V+ I+). The lowest levels were observed in non-infected, non-vaccinated participants (V-I-), while only infected (V-I+) or only vaccinated (V+ I-) individuals had intermediate levels. For the N protein, V+ I- and V-I+ participants exhibited high levels of Abs, while V+ I- and V-I-stayed low. **(d)** Most participants had similar levels of anti-EBNA1 Abs, with lower levels observed in some individuals spread across the immunological states, representing a minority of participants who have not encountered the Epstein-Barr virus. **(e)** Seroclusters resulting from hierarchical clustering of the anti-SARS-CoV-2 Abs. The clusters match self-reported immunological states well, with some mismatches where Ab levels deviate. **(f)** Individuals who self-reported only one dose of a vaccine were more likely to be reclassified to seroclusters representing non-vaccinated participants. **(g)** The time since vaccination also influences whether individuals with self-reported vaccination status were reclassified to seroclusters representing their vaccination status. **(h)** The number of months between infection and sampling showed waning levels of anti-N Abs in serocluster 4. **(i)** Serocluster 2 had stable Ab levels post-vaccination, where those for anti-S Ab levels were higher than for anti-N Ab. Smooth estimates from LOESS regression are displayed for anti-N Abs and anti-S Abs.

Our DBS serology survey confirms existing knowledge about seroconversion and points to the possibility of even differentiating between vaccination and natural infection.

### Seroclustering

To obtain even finer-grained insights into the actual antibody response at sampling and rely less on the self-reported infection and vaccination statuses, data-driven analysis of multi-analyte serology was used to stratify the samples. As shown in Fig 2e, hierarchical clustering revealed four main groups, denoted as seroclusters. Each serocluster mainly contained one of the four expected combinations of infection (I) and vaccination (V), which aligns well with our previous analysis. However, this classification allowed us to reduce discrepancies between reported events and measured immune response. Reasons for such discrepancies include asymptomatic infections, incorrect reports, time differences between sampling and events, or compromised immune responses.

As summarised in Table 3, serocluster 1 was the largest cluster (54% of all), with just 2% seropositive samples. Despite our serological results, 39 (16.5%) donors reported vaccination and 18 (7.6%) reported a previous SARS-CoV-2 infection. In Serocluster 2, > 90% of the donors self-reported to be vaccinated. Serology data supported this by revealing 95% S-positive and no N-positive samples. Serocluster 3 contained mostly those who reported an infection. This was supported by 97% being S+ and nearly >70% being N-. Serocluster 4 was the smallest cluster (8% of all), mainly containing participants who reported an infection and vaccination. These response phenotypes agreed well with the infection and vaccination statuses in the questionnaires.

**Table 3:** Seroclusters and self-reported demographics. Self-reported (apart from IFN AAb positivity) demographics within each serocluster. *P*-values are from Fisher exact tests.

| Seroclusters | 1 | 2 | 3 | 4 | P-value |
| --- | --- | --- | --- | --- | --- |
| **Participants** | 237 (54.2%) | 93 (21.3%) | 71 (16.3%) | 36 (8.2%) |  |
| **Serology** |  |  |  |  |  |
| N+ (% cluster) | 0 (0%) | 0 (0%) | 52 (73.2%) | 26 (72.2%) | < 0.001 |
| S+ (% cluster) | 5 (2.1%) | 88 (94.6%) | 69 (97.2%) | 36 (100%) | < 0.001 |
| **Age Group** |  |  |  |  |  |
| 18-29 | 54 (22.8%) | 6 (6.5%) | 14 (19.7%) | 2 (5.6%) | < 0.001 |
| 30-39 | 66 (27.8%) | 8 (8.6%) | 13 (18.3%) | 3 (8.3%) |  |
| 40-49 | 56 (23.6%) | 9 (9.7%) | 20 (28.2%) | 8 (22.2%) |  |
| 50-59 | 48 (20.3%) | 32 (34.4%) | 19 (26.8%) | 9 (25.0%) |  |
| 60-69 | 13 (5.5%) | 38 (40.9%) | 5 (7.0%) | 14 (38.9%) |  |
| **Sex** |  |  |  |  |  |
| Female | 131 (55.3%) | 68 (73.1%) | 40 (56.3%) | 21 (58.3%) | 0.013 |
| Male | 106 (44.7%) | 24 (25.8%) | 30 (42.3%) | 15 (41.7%) |  |
| Missing | 0 (0%) | 1 (1.1%) | 1 (1.4%) | 0 (0%) |  |
| **Region** |  |  |  |  |  |
| Gothenburg | 131 (55.3%) | 41 (44.1%) | 35 (49.3%) | 13 (36.1%) | 0.084 |
| Stockholm | 106 (44.7%) | 52 (55.9%) | 36 (50.7%) | 23 (63.9%) |  |
| **Symptoms (loss of)** |  |  |  |  |  |
| None | 218 (92.0%) | 82 (88.2%) | 32 (45.1%) | 16 (44.4%) | < 0.001 |
| Smell | 5 (2.1%) | 2 (2.2%) | 7 (9.9%) | 4 (11.1%) |  |
| Taste | 1 (0.4%) | 2 (2.2%) | 4 (5.6%) | 1 (2.8%) |  |
| Smell + Taste | 13 (5.5%) | 7 (7.5%) | 28 (39.4%) | 15 (41.7%) |  |
| **Vaccination** |  |  |  |  |  |
| Not vaccinated | 198 (83.5%) | 7 (7.5%) | 62 (87.3%) | 1 (2.8%) | < 0.001 |
| 1 dose | 39 (16.5%) | 64 (68.8%) | 8 (11.3%) | 22 (61.1%) |  |
| 2 doses | 0 (0%) | 22 (23.7%) | 0 (0%) | 13 (36.1%) |  |
| Missing | 0 (0%) | 0 (0%) | 1 (1.4%) | 0 (0%) |  |
| **Infection** |  |  |  |  |  |
| No | 219 (92.4%) | 85 (91.4%) | 15 (21.1%) | 4 (11.1%) | < 0.001 |
| Yes | 18 (7.6%) | 8 (8.6%) | 56 (78.9%) | 32 (88.9%) |  |
| **IFN AAb positivity** |  |  |  |  |  |
| No | 191 (80.6%) | 75 (80.6%) | 48 (67.6%) | 31 (86.1%) | 0.083 |
| Yes | 46 (19.4%) | 18 (19.4%) | 23 (32.4%) | 5 (13.9%) |  |

To quantify the general predictive power of anti-SARS-CoV-2 Abs and the improvement from introducing the seroclusters, the levels of anti-SARS-CoV-2 Abs were used to predict the questionnaire traits age, sex, region, infection, and vaccination, as well as seroclusters. We used a Lasso regularized multinomial regression, where 70% of the samples were used for training and 30% for testing. The penalty was tuned via five-fold cross-validation in the training set. The test set was used to evaluate the area under the curve (AUC) of the receiver operating curve (ROC) for each trait. Predictions of age, sex, and region were poor, with AUCs around 0.6 (Hand Till average for age), while the AUC was 0.85 for vaccination and 0.89 for infection. The average AUC for the prediction of serocluster was 0.99, ranging between 0.95 and 1.0 for the individual seroclusters (Fig S1, Table S2), showing that stratification of the response into multiple subgroups of vaccination and infection allows for better classification of the individuals.

In summary, data-driven seroclusters provided a finer-grained description of the molecular immune status at sampling. This reclassification may create more informed links between virus exposure, vaccination effect, seroconversion, and other molecular health-related variables.

### Mismatches between serocluster and questionnaire classifications

We observed noticeable differences between the molecular and self-reported data. Such discrepancies could contribute to the outcomes of connecting the classifications with other data. For example, we evaluated the seroclusters with the remaining questionnaire data. Seroclusters 1 and 3 contained more younger donors than seroclusters 2 and 4. Additionally, serocluster 2 contained a higher proportion of female donors than other seroclusters. No differences in the distribution of the regions were found, and self-reported symptoms matched the information on infections.

Still, about 11% of the samples (N = 47) were classified into seroclusters composed predominantly of non-vaccinated subjects despite self-reporting vaccination. To elucidate possible reasons, we compared the mismatch of donors with vaccinated participants who matched the expected classification. As shown in Fig. 2f-g, the discrepancy was due to the number of doses and the time between sampling and vaccine administration, but not the vaccine type (data not shown). This shows that seroclustering captured subtle differences in seroconversion, which is important when comparing COVID-related immune states.

### Effect of time since an event

To understand the contribution of time elapsed between COVID-19-related events and sampling in this cross-sectional study, we checked for the trends in antibody levels using the self-reported event periods. As shown in Fig 2h for participants from Serocluster 4, the levels of anti-S antibodies remained stable over 15 months while levels of anti-N decreased with time. In serocluster 2 on the other hand (Fig 2i), anti-S levels remained elevated, and anti-N levels remained low. This investigation demonstrated the influence of time since the event and that the anti-N and anti-S Ab levels behaved differently.

### Autoreactive antibodies against interferons

Autoreactive antibodies (AAbs) against interferons (IFN) have been described in COVID-

19 patients [53]. Hence, the occurrence and frequency of anti-IFN AAbs in the population-derived DBS samples could reveal relevant relationships between SARS-CoV-2 status, seroclusters, and disease severity. Utilizing the multiplexing capacity of the bead-based platform used for serology, we analyzed the IgG auto-reactivity against 22 full-length IFN proteins in the same assay as the COVID antigens. Similar to COVID-19 serology, the absence of a universal standard in DBS samples required us to classify AAb-positive samples at 12x SD above the IgG level population peak for each IFN. As shown in Table S1, we found circulating anti-IFN AAbs against all tested IFNs but IFN beta (IFNB), with frequencies ranging from 0.2% to 9% with a mean of 2.1% (95% CI: 1.2-3.0). AAbs against IFN alpha 17 (IFNA17) were most prevalent (8.9%), followed by IFN gamma (IFNG, 5.7%) and IFN omega (IFNW) 1 (4.8%). Anti-IFN AAbs were detected in 92 the donors (21% of all).

We then determined the frequencies of anti-IFN AAbs across the four seroclusters and noticed the highest frequencies in Serocluster 3, in which > 80% self-reported infections. This serocluster contained the highest percentage of samples (6.2%) classified as anti-IFN+, reaching frequencies of up to 9.9% for IFNA17, IFNA10 and IFNA8. The other seroclusters, containing uninfected or vaccinated subjects, had lower frequencies in anti-IFN AAbs. This suggests that recent natural infections rather than vaccinations influenced the occurrence of anti-IFN AAbs. To further understand these observations, we analyzed the AAb frequencies by sex, age, symptoms, and seropositivity. As detailed in Fig S2, AAb frequencies were higher in women (p<0.05) for IFN gamma (IFNG). Across age ranges, no AAb was found to differ in AAb frequencies (Fig S3). However, women below 40 years had a higher, albeit not statistically significant, prevalence of anti-IFNW1 (8.3%), anti-IFNA17 (14.4%), and anti-IFNG (10.3%) AAbs (Fig S6) compared to women above 40 years of age and men. As Fig S4 showed, frequencies of AAbs were linked to N-seropositivity across all IFNA subfamily members except IFNA4, IFNA16, and IFNA17. Differences in frequencies for S+ samples were comparable to N+ but with fewer significant differences (Fig S5). None of the highly prevalent AAbs (IFNA17, IFNG, and IFNW1) were linked to seropositivity. We did not find that elevated levels of IgG or IgM co-occurred with the traits or seroclusters. As shown in Fig 3, Fisher’s exact tests revealed a dominant association of AAbs against members of the IFN alpha subfamily in N+ samples. Such co-occurrence was less pronounced for S+ samples or self-reported symptoms. A reduced association for the S+ trait can be explained by most vaccinated subjects being S+ but not N+.

**Fig 3:**
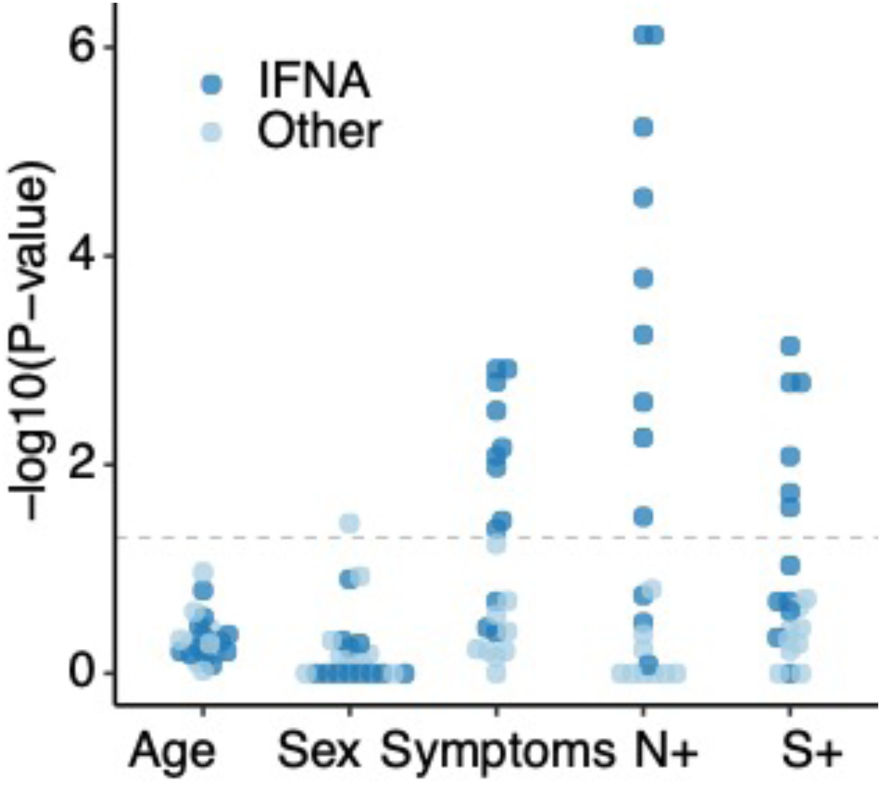
Differences in anti-IFN AAb frequencies across traits and proteins. Associations between interferon AAb positivity and age, sex, symptoms, and S and N seropositivity were tested using Fisher’s exact test. Each point represents the -log10(*P-value)* of a test for a trait and an interferon. The point is dark blue if the antigen is of the interferon A family. The grey dashed line marks a P-value of 0.05.

Our anti-IFN AAb data points to elevated reactivity levels of AAbs against the IFNA subfamily in primarily infected (N+) participants and in women under 40.

### Response phenotyping by the circulating proteome

The serological and autoantibody analyses assisted us in classifying donors based on their immune response to natural infections and vaccinations. To gain deeper insights into a wider range of molecular processes influenced by the exposures, we studied > 500 inflammation-related proteins in our DBS samples.

### Proteomics data

We used two complementary affinity proteomics methods built on dual antibody recognition and DNA-based readout. Each platform, branded to cover 368 and 250 targets, has established multiplexed assays to measure circulating proteins described to be involved in inflammation. Data from 395 common samples remained after data QC of each platform and ProtPQN normalisation [45] to account for layperson sampling.

Out of all 502 unique targets, 110 overlapped (Fig S7g), offering an immediate possibility to validate the results. As shown in Fig S7h, comparative analysis revealed 60 protein profiles to correlate between the two methods (rho > 0.5). A clear trend of higher correlations was found when both methods detected their target above the limit of detection (LOD) in >50% of the samples when using the supplier-provided LOD for plasma (Table S3). Both platforms delivered highly precise data, with 508 assays detected above LOD in >50% of the samples and CVs < 10% for 51% (median 9.9%) of the targets (Fig S7e). These 508 assays, targeting 435 unique proteins, were used for downstream analyses. Global analysis to identify outliers revealed two outlier samples, one of which was detected in both platforms (Fig S7a-b), while PCA did not reveal any imminent clusters that could have biased the downstream analyses (Fig S7c). Checking the protein data for differences in levels across all samples (Fig S7f), the widely expressed adaptor protein CRKL was the most stable protein. The monocyte protease CTSS, or cathepsin S, varied the most across the DBS samples. However, the bimodal distribution of protein contributed to the high variability. The second most variable protein was the cytokine IL36G which, according to the Human Protein Atlas [54], is expressed in respiratory epithelial cells, skin and tonsil.

Our findings confirm previous investigations with plasma samples [21], thus showing the utility of both methods to DBS analysis. Cross-platform comparisons are of growing interest in the community, especially when large sample sets and other omics data are available [55]. Being limited to self-reported information to benchmark our DBS proteomics assays, we included those proteins for which reported values were above LOD in more than 50% of all samples.

### Associations of circulating proteins with SARS-CoV-2 serology and IFN autoimmunity

We then used the data obtained by the two proteomics methods to determine the relationships of proteins with individual antigens of the humoral immune response to infection and related to auto-reactivity (Table S4). We limited this analysis to protein assays showing concordant nominal cross-platform associations.

For SARS-CoV-2, several immune-related proteins were associated with S and RBD. As shown in Fig 4, this includes positive associations with TREM2, a protein regulating the balance between anti-inflammatory and pro-inflammatory responses [56]. Similar trends were observed for the inflammation-promoting chemokine CXCL8 (IL8) and the immune-cell activating chemokine CXCL9 (MIG). However, a negative association was found between S/RBD and the acute phase, neutrophil-attracting chemokine CXCL1, suggesting that most samples were obtained after the acute phase of the SARS-CoV-2 infection. This aligns with the negative association found for the inhibitory receptor CD200R1, which has reduced levels despite higher levels of anti-SARS-CoV-2 Abs. There were less pronounced and platform-inconsistent associations between the circulating T-cell receptor TNFRSF4 (CD134) and CD200R1 with the N protein of the virus.

**Fig 4.**
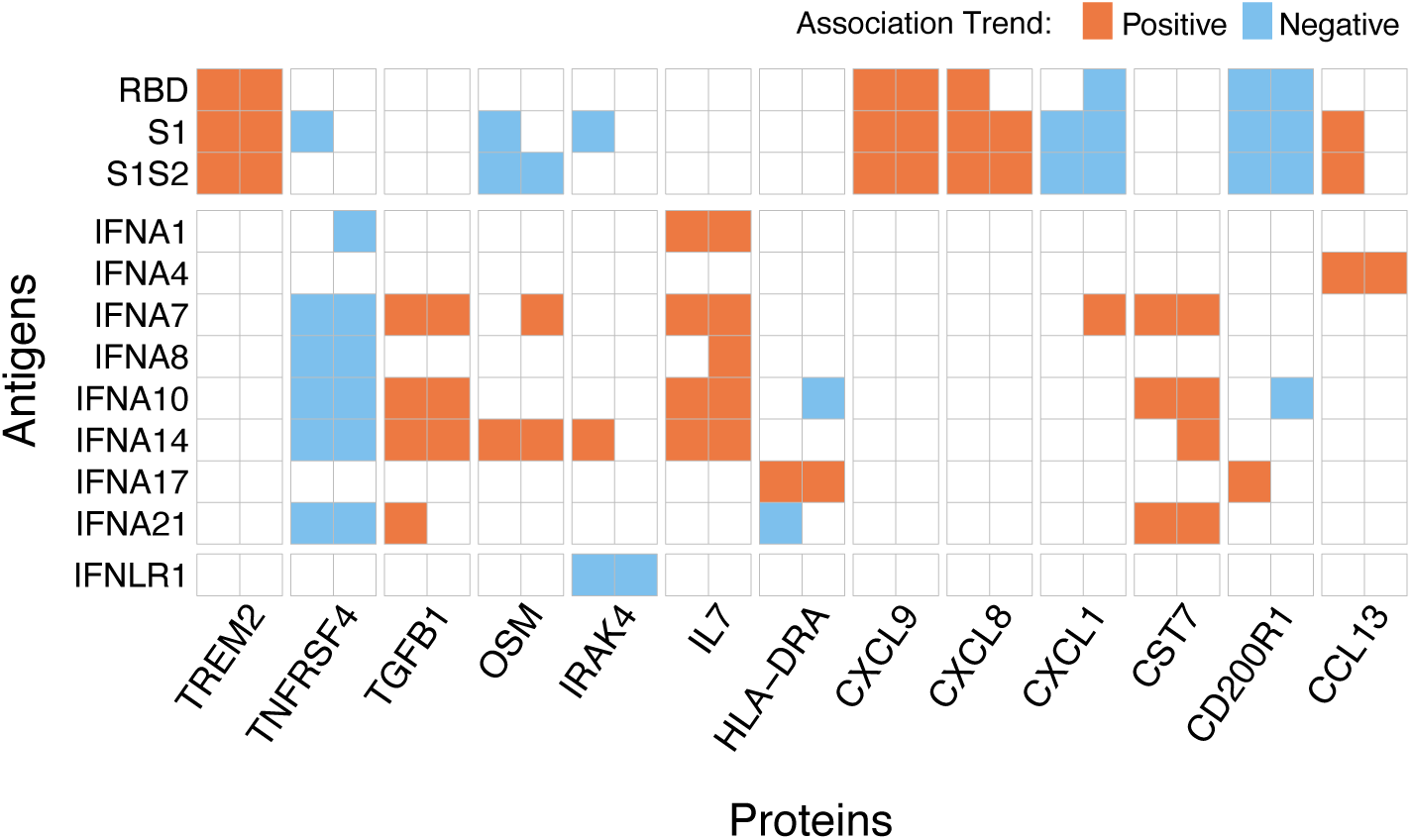
Relationships between circulating proteins and antibodies. Proteomics data from both platforms was used to associate circulating proteins with antibodies against both SARS-CoV-2 antigens and human interferons. From logistic regression analyses, only association pairs with concordant trends and nominal *P*-values < 0.05 are described. Red cells refer to positive association trends, meaning higher protein levels in the antibody-positive groups, while blue cells refer to negative associations.

For IFN AAbs, the association analysis with proteomics revealed eight proteins, most related to cytokine signaling. As shown in Fig 4, this included associations with more than one AAb against interferon alpha family members. For example, the protease inhibitor CST7, the T cell development cytokine IL7, and the regulatory growth factor TGFB1 had a positive association with several type I interferons. IFNA17, the target with the highest AAb frequency, was only associated with increasing levels of HLA-DRA, the alpha subunit of HLA-DR, an MHC class II receptor found on antigen-presenting cells. The T-cell activating receptor TNFRSF4 was negatively associated with anti-IFN AAbs. For other tested interferons, only negative associations of anti-IFNLR1 AAbs and IRAK4, a protease activating the innate immune response, were observed and platform consistent.

The analysis of circulating proteins in connection with antibodies against viral and human antigens allowed us to describe cellular processes linked to immune regulation and activation.

### Circulating proteins associated with seroclusters

The previously described seroclusters reflect the immune response against several viral antigens and present a more refined relationship between infection, vaccination, and their combination. Here, we investigated possible links between these response phenotypes and circulating proteins. There were 18 (3.5% of 508) proteins significantly associated (FDR < 0.05) with seroclusters (Fig 5, Table S5, S6). This included the above-mentioned CXCL9 and TREM2. Typically, seroclusters 2 and 4 had higher or lower protein levels than seroclusters 1 and 3. The endothelial leucine aminopeptidase 3 (LAP3), involved in protein degradation, had increased levels in seroclusters related to infection and/or vaccination. The protein HAVCR1, also called KIM-1, has been described to be involved in various biological processes, such as a host cell virus receptor, immune regulation, and kidney injury. It was elevated in seroclusters linked to vaccination. Inversely, the extracellular protein MEPE and the endocrine neuropeptide GAL decreased in a similar pattern for these seroclusters. Other noteworthy observations concerned differences between serocluster 2 (vaccinated but uninfected) and other seroclusters, reflecting different molecular processes. For example, lower levels were found for AGRP, an endocrine hormone in food and energy control; CRLF1, a cytokine secreted by smooth muscles, different fibroblasts, and brain cells for cell growth and differentiation; and SCG3, a protein secreted by brain cells and endocrine tissue to form secretory granules. Higher levels in serocluster 2 were found for CXCL9, the gastrointestinal regulator protein MLN, and CXCL17, a chemokine secreted by salivary and respiratory epithelium and glands in the stomach.

**Fig 5.**
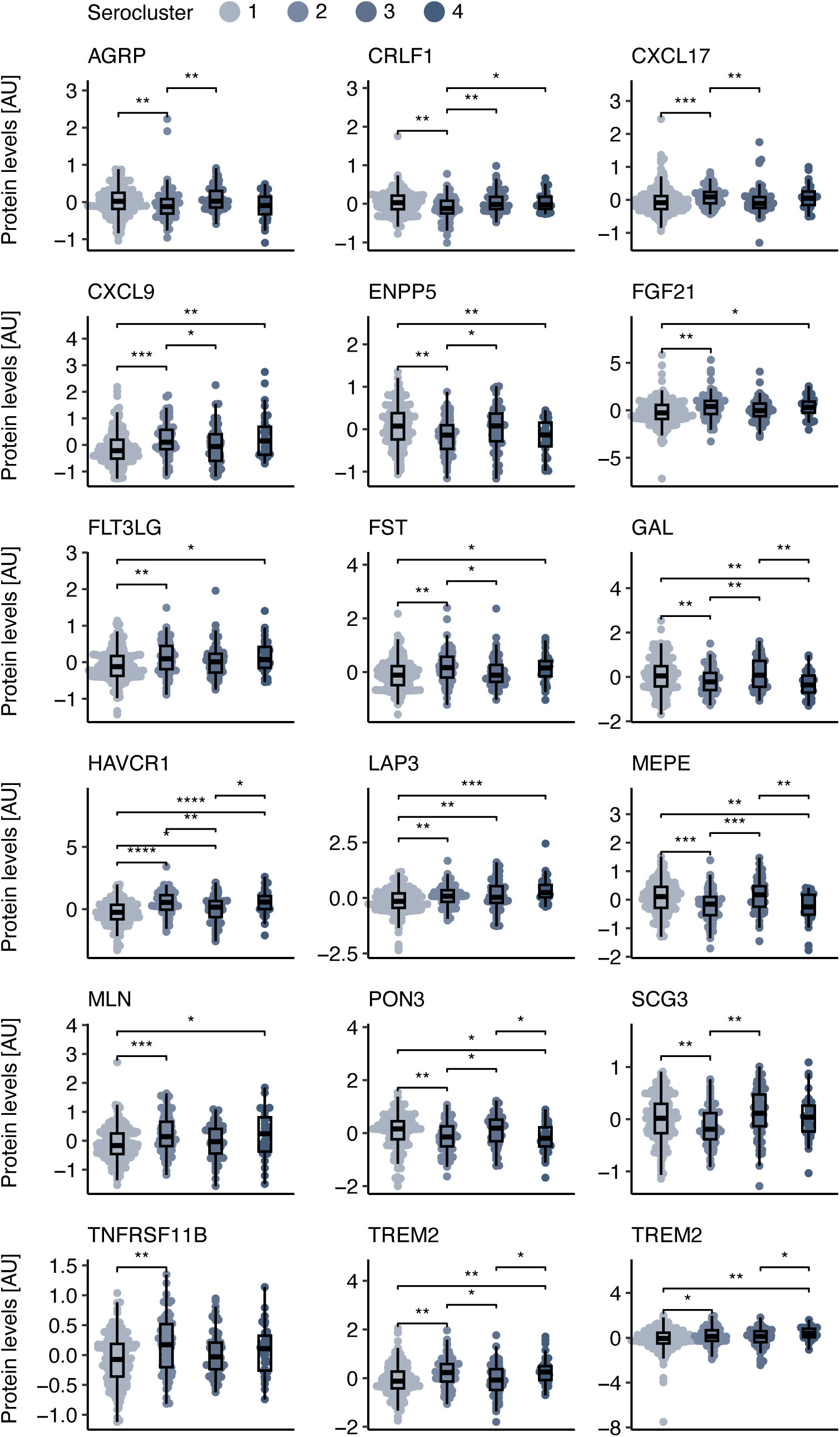
Linear associations between seroclusters and proteins. Proteins with significantly different levels between the seroclusters are shown. Seroclusters are depicted in shades of blue. Asterisks denote the degree of significance from Kruskal-Wallis tests with *: FDR<0.05, **: FDR<0.01, ***: FDR<0.001, and ****: FDR<0.0001. The center line in the boxes represents the median value and whiskers the upper and lower 25%.

Identifying proteins from different physiological processes suggests that traits other than COVID-related proteins may drive the differences between the seroclusters. Age, sex, BMI, genetics, lifestyle, liver and kidney function, smoking or medication use are well-established factors that may influence the circulating proteome and confound disease-specific investigations [49]. In our investigation we tested for age and sex associations (Fig S8, Table S7) as well as COVID-related traits. The latter investigations point to plausible links between seropositivity and individual immune response. Due to the vaccine rollout strategy in Sweden, there is an intrinsic link between age and vaccination.

### Circulating proteins for predicting traits

To evaluate the data, we performed a multivariate Lasso regression analysis, similarly to the serology data, to predict different traits with circulating proteins (Fig 6, Table S8).

**Fig 6.**
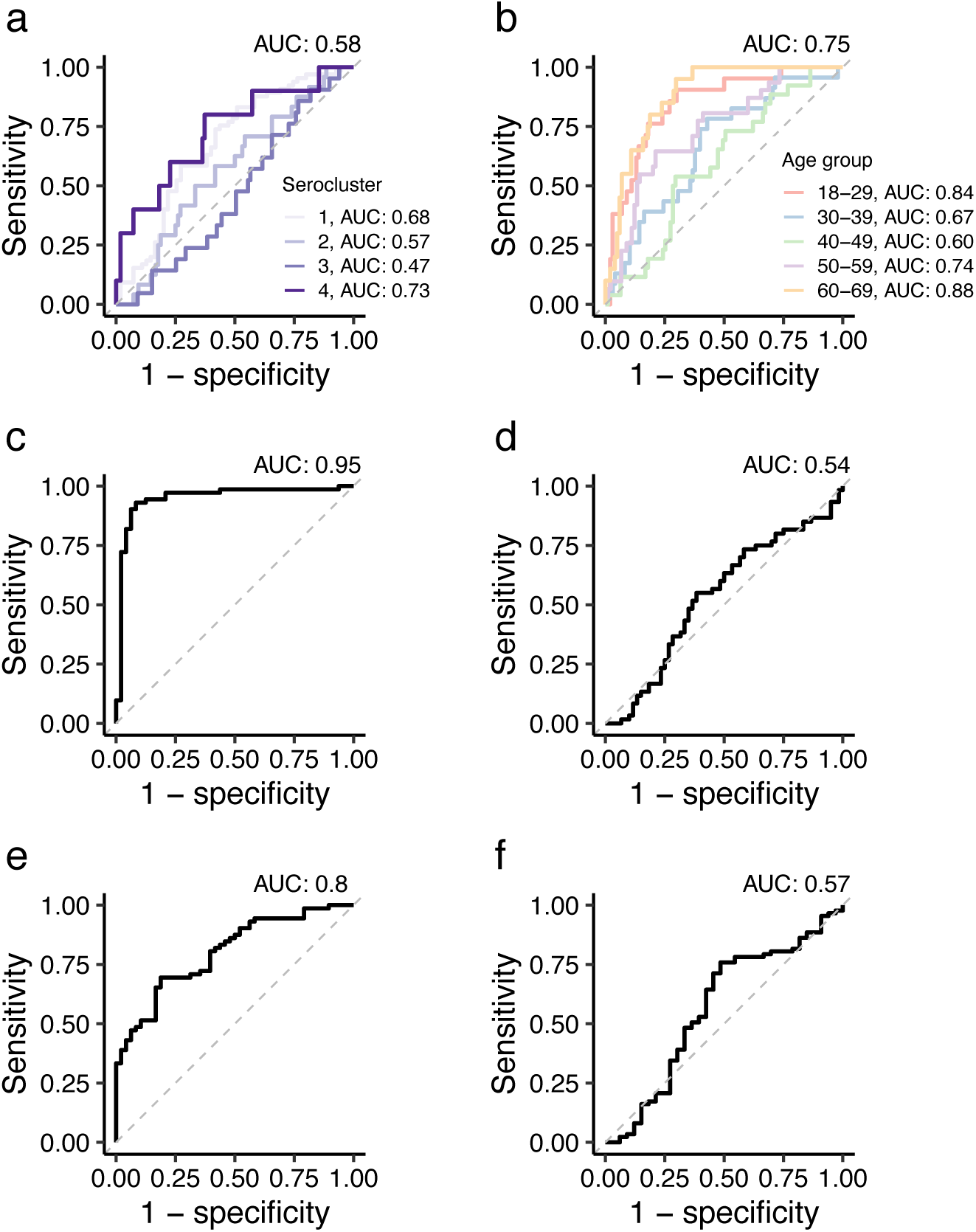
Predicting traits using circulating proteins. ROC curves for prediction of **(a)** serocluster, (**b**) age, (**c**) sex, (**d**) region, (**e**) vaccination, and (**f**) infection using circulating proteins as predictors in penalized multinomial regression. For serocluster and age prediction with more than two classes, the AUC shown at the top of the plot is the average AUC from the Hand Till method.

Protein-based prediction performed best for vaccinated and infected groups while remaining poorer predictors for other seroclusters (Fig 6a; AUC 0.47 to 0.73). Proteins were also better predictors for the youngest and oldest age groups than the intermediate age groups (Fig 6b; AUC 0.60 to 0.88). Reassuringly, and given the high importance of sex for human physiology, our circulating proteome data contained sufficient sex-specific information to predict this trait (Fig 6c; AUC 0.95). In addition, the low AUC for predicting sampling region suggests that the geographical location had a limited effect on the proteome when sampling follows a unified protocol (Fig 6d; AUC 0.54). The proteomics data was a weak predictor for self-reported infection (Fig 6e; AUC 0.57), which reflects the low number of acute infections among many that have already passed. Self-reported vaccination could be predicted moderately well (Fig 6f; AUC 0.80). This can be attributed to the recent administration events but also reflects a bias towards elderly individuals, who were prioritized. After adjusting the protein levels for age, the AUC decreased to 0.5.

The circulating proteins allowed us to predict sex and some age groups at higher performance (AUC > 0.8) than the serology data (AUC < 0.7). As expected, serology outperformed proteomics for predicting previous infections, vaccinations, and antibody-defined seroclusters (AUC > 0.85).

### Proteomics-centric analysis

Thus far, our analyses have been based on self-reported questionnaire data and serological measurements, revealing proteins associated with different traits and seroclusters. We also observed that proteins might reflect other, possibly COVID-unrelated processes and that these influenced our investigations. Instead of relying on these traits or groupings, we chose a proteomics-centric, data-driven approach to identify health phenotypes.

The data from all 500 circulating proteins were categorized using Weighted Gene Correlation Network Analysis (WGCNA). This was chosen to reduce the number of variables originating from the 500 protein assays to fewer dimensions comparable to the serological analyses. This resulted in five distinct WGCNA modules, denoted by the colours blue (containing 69 proteins from 76 assays), brown (61 proteins, 70 assays), green (30 proteins, 32 assay), turquoise (72 proteins, 83 assays), and yellow (55 proteins, 65 assays), and a sixth module (grey) containing the remaining uncorrelated proteins (167 proteins, 182 assays) (Fig S9a-b, Table S9). Of 73 proteins measured with two methods, 54 (74%) had both assays assigned to the same module (Fig S9c).

To understand the underlying biology driving the formation of the WGCNA modules, we related the modules to previous studies of protein associations with 28 traits in the UK Biobank [49]. The proportions of proteins significantly associated with each trait were compared between each module and the remaining modules using the Fisher exact test (Fig S10). This showed that the blue module was enriched in proteins associated with smoking (MMP10), alveolar macrophages (AGRP), lung-specific proteins like SCGB1A1 (also called CC16), and kidney and liver function. The brown module was rich in proteins associated with age and many other health-related aspects tested in the UKB study. Inversely, the green and yellow modules contained fewer proteins associated with sex and health phenotypes, including smoking and lower respiratory infection. The turquoise module was enriched in proteins associated with acute lower respiratory infection.

To link the modules further to immune function, we investigated the immune cell enrichment of proteins in each module (Fig S11). Proteins from the turquoise module were enriched in cells such as neutrophils, eosinophils, and dendritic cells while not in T cells, suggesting that the module is connected to the innate immune system. The STRING database (version 12.0) was used to study the functional enrichment of each module (Fig S12). The blue module was linked to T-cell function, the brown, turquoise and grey modules to migration, chemotaxis and cytokine response, and the yellow module to intracellular signalling through pathways such as the mitogen-activated protein kinase (MAPK) and phosphoinositide 3-kinase (PI3K) pathways. The green module was linked to cell signalling (MAPK) and immune response (NF-kappa B), as well as recognition of pathogen DNA (via Toll-like receptor 9), suggesting a role in recent infection by pathogens other than SARS-CoV-2.

From the five distinct WGCNA modules, we computed the module eigengenes (MEs), which are the first principal components of the constituent proteins. These five MEs were then used for clustering, revealing 14 protein-based phenotypes, denoted proteotypes. Stability analysis using the mean Jaccard index (MJI) shortlisted five stable clusters which met the criteria of containing N > 10 individuals and an MJI > 0.5. The five proteotypes represented 24% (N = 95) of all donors and were labeled (1-5) in order of decreasing Ns of 30, 20, 18, 16, and 11, respectively. The remaining less stable proteotypes were aggregated into proteotype 0, containing 76% of all participants (N = 300).

The proteotypes were visualised in a PCA biplot of the MEs (Fig 7a), showing that the five proteotypes cluster around the perimeter of the PC1-PC2 space, but less so for PC3 and PC4 (Fig S13). In Fig 7b, the distinct WGCNA module profiles of proteotypes 1-5 were compared to the other proteotypes (including 0). Effect sizes were used to standardize the differences in relative protein levels between the phenotypes. For proteotype 1, effect sizes were negative across all modules. Proteotype 2 showed positive effect sizes for the turquoise module, while those for the blue and brown modules remained negative. Proteotype 3 was characterised by positive effect sizes across all modules. Effect sizes of proteotype 4 were positive for the blue and brown modules and negative for all others. Proteotype 5 showed the largest positive effect sizes for the blue and brown modules and the grey module. This analysis also showed that some proteotypes shared trends and magnitudes of effect sizes. For example, proteotypes 3 and 4 were similar for the blue and brown modules but different for the green, turquoise, and yellow ones.

**Fig 7.**
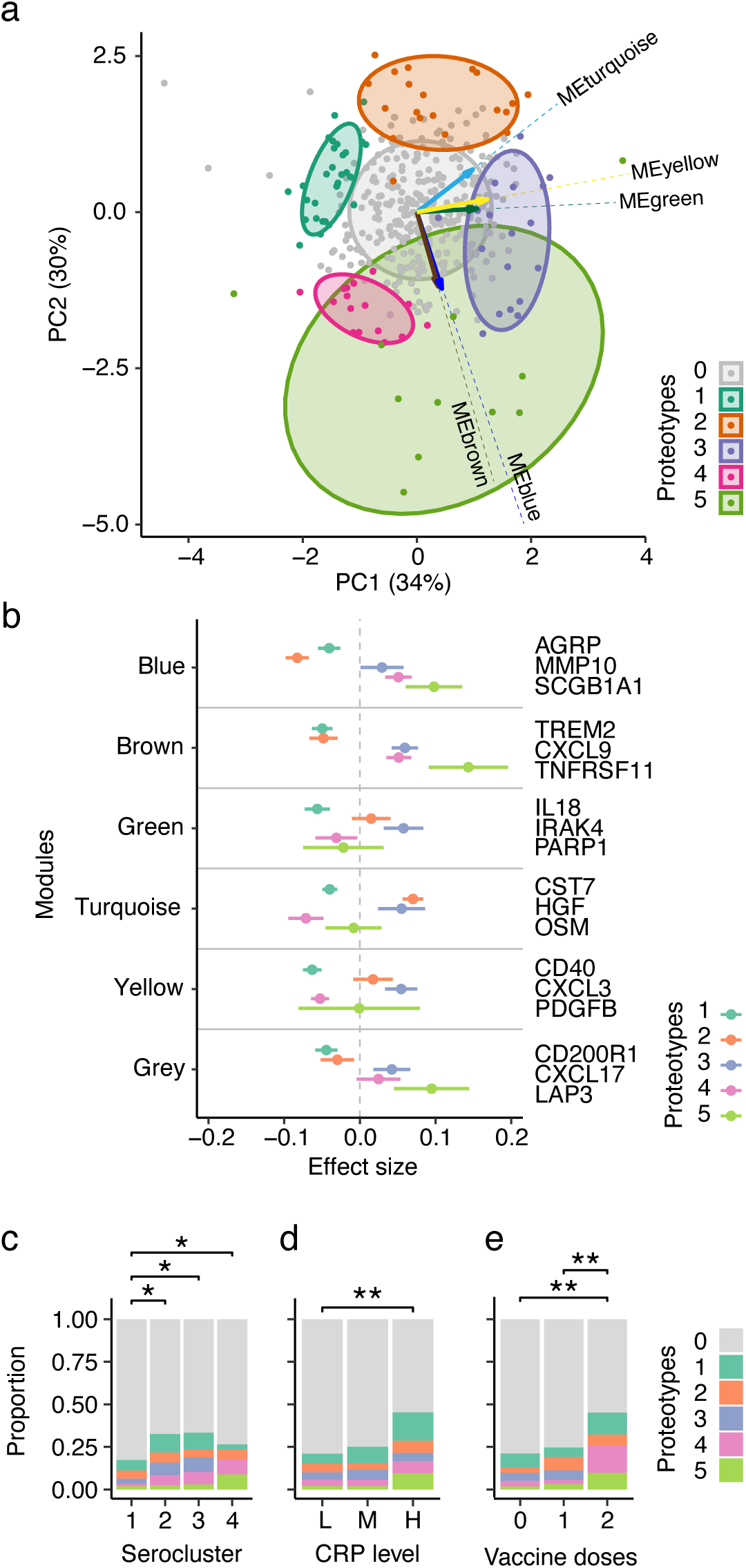
Clustering of samples based on WGCNA protein modules. **(a)** PCA biplot of the WGCNA MEs, coloured by proteotype membership. **(b)** Comparison of module effect sizes of the proteotype. The effect sizes were measured as the difference in ME mean between each proteotype 1-5 and the remaining proteotypes. The horizontal bars show the 95% confidence intervals. Three examples of proteins from each module are shown to the right. **(c)** Distributions of proteotypes within each serocluster. **(d)** Distributions of proteotypes across low (L), medium (M), and high (H) CRP levels. **(e)** Distributions of proteotypes across the number of vaccine doses. Asterisks denote the Fisher exact test *P*-*value* range (*: *P*<0.05, **: *P*<0.01).

To further classify the proteotypes, we checked for differences in questionnaire information (Table 4). In contrast to serology, there were no significant differences in age, sex, or self-reported symptoms. The influence of infection, vaccination, and seroclustering remained observable but was more evenly distributed across the phenotype. Still, seronegative individuals from serocluster 1 had a lower proteotype heterogeneity than the other seropositive clusters (Fig 7c), pointing to the influence of infections and vaccinations on the proteomic profiles. Notably, anti-IFN AAbs were less frequent in proteotype 2 (5%), consisting of mostly uninfected donors, and most prevalent in proteotype 3 (28%), although not statistically significant (Fisher exact test *P* = 0.5). The occurrence of IFN-AAbs was independent of CRP levels (Fisher exact test *P* = 0.6).

**Table 4:** Demographics of proteomics phenotype. Demographics within each proteotype. All but serocluster, CRP level, and IFN AAb positivity are self-reported. *P*-values are from Fisher exact tests.

| Proteotypes | 0 | 1 | 2 | 3 | 4 | 5 | P-value |
| --- | --- | --- | --- | --- | --- | --- | --- |
| **Participants** | 300 (75.9%) | 30 (7.6%) | 20 (5.1%) | 18 (4.6%) | 16 (4.1%) | 11 (2.8%) |  |
| **Age group** |  |  |  |  |  |  |  |
| 18-29 | 52 (17.3%) | 8 (26.7%) | 3 (15.0%) | 5 (27.8%) | 2 (12.5%) | 0 (0%) | 0.24 |
| 30-39 | 60 (20.0%) | 3 (10.0%) | 3 (15.0%) | 3 (16.7%) | 3 (18.8%) | 4 (36.4%) |  |
| 40-49 | 64 (21.3%) | 9 (30.0%) | 7 (35.0%) | 1 (5.6%) | 3 (18.8%) | 0 (0%) |  |
| 50-59 | 80 (26.7%) | 5 (16.7%) | 3 (15.0%) | 6 (33.3%) | 3 (18.8%) | 3 (27.3%) |  |
| 60-69 | 44 (14.7%) | 5 (16.7%) | 4 (20.0%) | 3 (16.7%) | 5 (31.3%) | 4 (36.4%) |  |
| **Sex** |  |  |  |  |  |  |  |
| Female | 183 (61.0%) | 17 (56.7%) | 10 (50.0%) | 10 (55.6%) | 11 (68.8%) | 6 (54.5%) | 0.86 |
| Male | 117 (39.0%) | 13 (43.3%) | 10 (50.0%) | 8 (44.4%) | 5 (31.3%) | 5 (45.5%) |  |
| **Infection status** |  |  |  |  |  |  |  |
| 0 | 225 (75.0%) | 22 (73.3%) | 16 (80.0%) | 12 (66.7%) | 8 (50.0%) | 4 (36.4%) | 0.025 |
| 1 | 75 (25.0%) | 8 (26.7%) | 4 (20.0%) | 6 (33.3%) | 8 (50.0%) | 7 (63.6%) |  |
| **Vaccination status** |  |  |  |  |  |  |  |
| 0 | 187 (62.3%) | 19 (63.3%) | 8 (40.0%) | 11 (61.1%) | 8 (50.0%) | 4 (36.4%) | 0.005 |
| 1 | 95 (31.7%) | 7 (23.3%) | 10 (50.0%) | 7 (38.9%) | 3 (18.8%) | 4 (36.4%) |  |
| 2 | 17 (5.7%) | 4 (13.3%) | 2 (10.0%) | 0 (0%) | 5 (31.3%) | 3 (27.3%) |  |
| Missing | 1 (0.3%) | 0 (0%) | 0 (0%) | 0 (0%) | 0 (0%) | 0 (0%) |  |
| **Symptoms (loss of)** |  |  |  |  |  |  |  |
| None | 245 (81.7%) | 25 (83.3%) | 15 (75.0%) | 14 (77.8%) | 10 (62.5%) | 6 (54.5%) | 0.14 |
| Smell | 8 (2.7%) | 1 (3.3%) | 2 (10.0%) | 2 (11.1%) | 1 (6.3%) | 1 (9.1%) |  |
| Taste | 6 (2.0%) | 0 (0%) | 0 (0%) | 0 (0%) | 0 (0%) | 1 (9.1%) |  |
| Smell + Taste | 41 (13.7%) | 4 (13.3%) | 3 (15.0%) | 2 (11.1%) | 5 (31.3%) | 3 (27.3%) |  |
| **Serocluster** |  |  |  |  |  |  |  |
| 1 | 172 (57.3%) | 13 (43.3%) | 10 (50.0%) | 6 (33.3%) | 3 (18.8%) | 4 (36.4%) | 0.025 |
| 2 | 56 (18.7%) | 9 (30.0%) | 5 (25.0%) | 6 (33.3%) | 5 (31.3%) | 2 (18.2%) |  |
| 3 | 47 (15.7%) | 7 (23.3%) | 3 (15.0%) | 6 (33.3%) | 5 (31.3%) | 2 (18.2%) |  |
| 4 | 25 (8.3%) | 1 (3.3%) | 2 (10.0%) | 0 (0%) | 3 (18.8%) | 3 (27.3%) |  |
| **CRP level category** |  |  |  |  |  |  |  |
| Low | 238 (79.3%) | 18 (60.0%) | 15 (75.0%) | 13 (72.2%) | 11 (68.8%) | 6 (54.5%) | 0.045 |
| Medium | 39 (13.0%) | 5 (16.7%) | 2 (10.0%) | 3 (16.7%) | 2 (12.5%) | 1 (9.1%) |  |
| High | 23 (7.7%) | 7 (23.3%) | 3 (15.0%) | 2 (11.1%) | 3 (18.8%) | 4 (36.4%) |  |
| **IFN AAb positivity** |  |  |  |  |  |  |  |
| No | 237 (79.0%) | 23 (76.7%) | 19 (95.0%) | 13 (72.2%) | 12 (75.0%) | 9 (81.8%) | 0.5 |
| Yes | 63 (21.0%) | 7 (23.3%) | 1 (5.0%) | 5 (27.8%) | 4 (25.0%) | 2 (18.2%) |  |

To further understand the seropositivity profiles of the proteotypes, we categorised the samples into high, medium, and low levels of the well-established but unspecific inflammation marker CRP (Fig 7d) based on 3 and 6 SD population-based thresholds. Proteotypes 1 and 5 contained high proportions of individuals with elevated CRP levels, indicating likely ongoing infection or inflammation. In contrast, proteotype 4 had high proportions of infection and vaccination, as indicated by the relatively low proportion of samples being assigned to serocluster 1, without elevated CRP levels, suggesting that the proteomic assays captured the time-sensitive immune response, complementing the serological assays (Fig 7e).

## Discussion

We performed serological and proteomic analyses on home-sampled dried blood in a general population to elucidate the features of immune response against natural infection and vaccination. Our study expands the suitability of self-sampling at home as an alternative for multi-molecular analysis comparable to studies using venous blood draws [6], [7], [21]. Others focused on clinical or informed cohort studies, but little remained known about the effects in the general population. By employing easy-to-perform quantitative sampling kits for random individuals, a reduced bias compared to self-enrollment studies was achieved while maintaining a low sampling failure rate in an untrained population. The timing of sampling, scheduled during a time when only a subset of individuals had been infected by or vaccinated against COVID-19, makes our study set unique. Due to the global spread of the virus and vaccines, similar sample sets remain difficult to be collected.

At first, the serological study revealed the expected immune biology and allowed us to infer the infection and vaccination status of individuals by the differences in immune response, which has been shown previously [57]. The assays only measured levels of IgG, meaning that we lack the insights that IgM levels could add to further disentangle the time course of disease response phenotypes. The multi-analyte data-driven seroclustering provided time-resolved molecular insights into the humoral response, allowing us, with high predictive precision (AUC > 0.99), to classify the individuals by their circulating antibody response. We also found differences between the reported and measured immune profiles, caused by the waning of anti-N antibodies over time and the incomplete development anti-S response due to recent vaccinations.

The multiplexed capacity of the bead array platform allowed us to study auto-reactive IgG against human interferons alongside the SARS-CoV-2 antigen. We found an enriched co-occurrence of anti-IFN AAbs in infected donors. Earlier discussions about anti-SARS-CoV-2 antibodies suggested that cross-reactivity may occur in vivo and cause disease pathologies [58], however, it remains controversial whether AAbs to IFNs existed prior to or only after infection. Interestingly, a recent study supports our observations as it suggested that mucosal antibodies against IFNs can appear temporarily even in milder cases [59]. Despite the need for further investigations with appropriate designs, it is reasonable to suggest that we could detect anti-IFN AAbs in DBS due to the presence of hematopoietic cells or interstitial fluid (in addition to the liquid plasma fraction). We cannot, however, exclude the possibility that anti-N antibodies cross-reacted with the immobilized type 1 interferons in our assay. In addition, we studied nine IFN proteins alongside their AAbs, however, there was no association between the two data types. Hence, our observations can add some insights to the sparse literature and an incomplete understanding of the cause and consequences of anti-IFN AAbs in milder cases of COVID-19.

It is well established that profiling of the circulating proteome has given important insights into acute and severe COVID-19 [60], and studies continue to emerge to gain more knowledge about longer-term consequences, such as post-COVID-19 sequelae. We chose and compared two complementary affinity proteomics platforms [23], [24] to determine immune-related proteins in dried blood spots. This expands previous work [21] into lower abundance proteins and towards more acute and shorter-lived signatures. The platform comparison matches with observations made in plasma samples [24], [61], highlighting which low abundant protein profiles were concordant and reliable, but also revealing discordances that require further investigations. Reassuringly, > 60% of the common proteins were confirmed by the other platform. Ultimately, the discrepancy between methods calls for further investigation with orthogonal methods to increase confidence in technology-dependent observations [8].

Adding proteomics to the serological and AAb data provided more fine-grained insights into physiological processes. The general low levels of well-known acute-phase proteins, such as CRP, IL6, TNF, or IFNG, suggested that most samples were taken after the acute phase of the infection. Other proteins provided information about non-COVID-19 related events (e.g., other respiratory infections or diseases). We did not collect information about other underlying chronic or acute diseases and could only infer this from the protein data collected at sampling. Hence, we do not know if and for how long proteins remained elevated compared to the pre-infection phase.

The stability of the molecular features appeared as a major differentiator of circulating proteins and anti-SARS-CoV-2 Abs. Since such anti-COVID Abs did not exist before infection, they must have been acquired during the pandemic. Certain circulating proteins, however, are known to be also stable and person-specific [62], [63], hence possible differentiators before the pandemic sampling. Still, other proteins may have changed in response to infection or vaccination [64], which could have made the profiles of some donor more similar or different. As such, each data type can provide a different view on the human immune response: anti-SARS-CoV-2 Abs are limited to COVID-19; anti-IFN AAbs could be acquired or pre-existing and relate to disease vulnerability or autoimmune conditions; while the pleiotropy of circulating proteins can reveal immune-related as well as a wider range of “unrelated” phenotypes. This is also reflected in using binary categories for antibody data compared to continuous scales and bidirectional changes for the circulating proteins.

To reduce the proteomics data to informative dimensions, we chose to compute WGCNA modules. Select sets of proteins point to different plausible phenotypes and provide clinically informative health projections, and insights into cell-mediated immune processes. Many of the module proteins were involved in COVID-19 pathologies: respiratory distress was revealed by MMP10 [65], found in respiratory epithelial cells; the lung-specific SCGB1A1 [66]; interactions with viruses through renal receptor HAVCR1 [67]; inflammation via CRP or pro-inflammatory chemokines like CXCL9 or immune cell receptors like TREM2 [68]. Our study also included other pleiotropic inflammation markers like IL6, TNF, and IFNG, which were shown to be elevated in acute disease when studying plasma samples [9], [69]. However, these were not shortlisted as informative features in our modules. We attribute this to the lower number of acute or active cases. Compared to stable antibody levels, a reduction in inflammation related proteins occurs after recovery and being symptom-free. We can, however, not exclude that the analytical performance of the assays in DBS samples was lower than in plasma.

Our study revealed subtle proteome differences between seroclusters, suggesting that traits other than the COVID-19-related traits or exposures influenced the phenotypes. This was observed in the age-related effects that increase the risk of diseases. In addition, our observations focused on the immune system, hence we evaluated how other valuable processes, such as coagulation or organ damage, influenced the molecular phenotypes. Still, the observations made in the circulating sub-proteome finds support in the literature where plasma from venous blood draws was used [69], [70], [71]. As discussed previously [21], the use of dried samples for multi-molecular analysis has just started. Hence, analytical factors, such as stability and structural integrity of dried-up proteins or leakage of intracellular proteins during the drying process, can influence the assay performance of any analyte, including those postulated by other studies.

We also acknowledge that our study has limitations, some of which are shared with other population proteomics efforts [72]. Compared to a previous study, which used samples from the first wave [6], fewer members of the general public participated (20% vs. 55%). This can be attributed to a wider range of COVID-19 countermeasures in Sweden, such as PCR testing, serology, and vaccination programs. Our study appears underpowered compared to other large-scale population proteomics efforts, like in the UK Biobank [49]. Still, coordinated sampling of random individuals from two geographically distant locations remains unique and limits collider bias during enrollment. Replicating our observations in other studies is possible but requires a careful matching of sampling year, seasonality, and urban environments with comparable pollution. This should include the impact of the COVID-19 situation and applied countermeasures, as well as lifestyle during the warmer months of the year. Nonetheless, we cannot exclude that previous infections, other chronic diseases or health conditions, unknown socioeconomic factors, misinterpretation of the questions, and the correctness of the provided information influence the participation rate. As we chose to keep the donors anonymous, we cannot recontact them to follow up longitudinally and provide missing health-related information. The donors’ overall health status, BMI, genetics, or medication use is currently still needed to define the clinical representations of the five proteotypes.

This study highlights the importance of understanding population-level immune responses to SARS-CoV-2 in the context of underlying health conditions. These may, depending on age, sex, socio-economic and many other factors, affect a large proportion of the population. Taking smoking as an example, 5% of Swedish adults report smoking daily and 6% report smoking occasionally [73]. The effect of this could be observed in our data with the blue protein module containing a high proportion of proteins related to smoking as well as proteins, such as MMP10 and SCGB1A1, related to respiratory distress. By identifying differences in immune responses and proteomes, these findings support the development of scalable tools to monitor and manage public health trajectories and resources effectively. These could also benefit studies of long-COVID since several of the presented markers, like CRP, CXCL9, anti-IFN AAbs and CST7, have been shortlisted [74]. Additionally, the use of minimally invasive methods like DBS self-sampling can enhance outreach to diverse populations and ensure equitable health surveillance.

In conclusion, our population survey provided insights into the molecular response to the SARS-CoV-2 virus and administered vaccines. The 25% of participants with a deviating health phenotype may serve a useful estimate when conducting similar surveys. It shows the heterogeneity of immune status when sampling random participants, the influence of sampling timing, and that other unrelated events may influence the assignment of molecular data to health events.

## Data and code availability

Data supporting the findings of this study will be deposited under the manuscript’s title at SciLifeLab Data Repository (https://scilifelab.figshare.com). The datasets are under restricted access since they represent individual-level human data. As described in the repository, access can be granted for non-commercial validation purposes upon reasonable request to the corresponding authors. Any unique or newly developed analysis code will be available upon publication at https://github.com/Schwenk-Lab.

## Acknowledgements

First, we thank all anonymous volunteers who provided blood samples and answered the questionnaire for our study. We thank Matilda Dale for her support and the members of the Schwenk and Nilsson Labs for fruitful discussions. We thank Benjamin Murrell (Karolinska Institutet) for providing help and code for normalising the serology data, and August Jernbom Falk for comments on the manuscript. We acknowledge the tremendous support from the SciLifeLab infrastructure units for Affinity Proteomics in Stockholm and Uppsala, and the Autoimmunity and Serology Profiling unit. We thank the KTH node of Protein Production Sweden (PPS), a national research infrastructure funded by the Swedish Research Council), the Human Secretome Project at the Wallenberg Center for Protein Research, and everyone at the Human Protein Atlas (HPA). We thank the teams at Olink Proteomics and Alamar Bioscience for their support.

## Author contributions

L.D., A.B., M.B.A., and V.A. curated data and performed data analysis. A.B. and H.A. performed experiments. M.K., A.M., and C.F. managed infrastructures and provided technical input. S. Bauer. provided input on machine learning and data analysis. E.M., S. Björkander and S.K.M. provided input on immune phenotypes and population studies. O.B., Å.T.N., M.G., N.R., and J.M.S. conceptualized the study and collected clinical specimens. J.M.S. and N.R. designed the experimental analyses and supervised the work. L.D. and J.M.S. wrote the original draft of the manuscript. All authors reviewed and edited the manuscript.

